# Development of an Inventory to Identify Psychosocial Factors Influencing Hand Usage: the CHUC

**DOI:** 10.64898/2026.03.26.26347326

**Authors:** Téa Soberano, Chih-Hung Chang, Alexandre J Marcori, Benjamin A Philip

## Abstract

**Objective:** To develop the first inventory to measure psychosocial concerns about use of the non-preferred hand, toward the long-term goal of identifying the casual factors of left-right hand choices (“hand usage”).

**Design:** Cross-sectional

**Setting:** Online question battery

**Participants:** 181 healthy adults

**Interventions:** Not applicable

**Main Outcome Measures:** Self-reported concerns about emotional and physical consequences of using the non-preferred hand.

**Results:** Emotional and physical consequences reflected internally consistent categories (Cronbach’s α > 0.9) that were moderately correlated with each other (ρ = 0.783 p = 0.002). Concerns were activity-dependent in each category (p < 1×10^-100^). Reliability analysis and principal components analysis were used to reduce the battery to the 51-item Changed Hand Usage Concerns inventory, which encompasses everyday tasks and concerns about physical and emotional consequences of using the non-preferred hand in those tasks.

**Conclusions:** Concerns about emotional vs. physical consequences of non-preferred hand use reflect coherent and internally consistent categories The Changed Hand Usage Concerns inventory allows assessment of psychosocial concerns about usage of the non-preferred hand for future attempts to manipulate hand usage via rehabilitation in patients with unilateral or asymmetric impairment.

## 1. INTRODUCTION

Handedness is an ancient and fundamental human trait [1], but little is known about how humans choose which hand to use for daily activities. Hand choices during action (also known as hand usage) are separable from performance asymmetries between the hands [2,3]. Here we focus on hand preferences during action, sometimes called hand choice or hand usage, in part because hand depends in part on psychosocial factors (for definition, see Engel, 1977) such as psychological factors and individual traits (e.g. anxiety, repression) [5,6], and the influence of social pressures during development [7,8]. Psychosocial influences on hand usage may be especially relevant in individuals with conditions that limit the use of the PH, as both social and psychological burdens can be enhanced after impairment [9].

Hand usage can provide an important lens for rehabilitation, especially in patients with unilateral impairment of one upper limb. In patients with unilateral peripheral nerve injury, decreased use of the affected hand is associated with poor health-related quality of life, activity participation, and disability; regardless of whether the affected hand is the PH or NPH [10]. Conversely, some patients may benefit from increased use of their NPH, especially after impairment to the PH makes the NPH their more-dexterous hand [11]. However, current rehabilitation and training methods have shown limited ability to alter hand usage [12–15]. This inability to alter hand usage arises, in part, because it remains unclear what an intervention should target beyond the motor domains, and whether psychosocial factors associated with emotional states and performance expectations may impede handedness-related rehabilitation.

Hand usage (left/right hand choices in an unconstrained context) is driven by a complex array of factors. Hand usage depends in part on performance asymmetries between the hands [16,17], but also on task-specific demands [18–20] and the reinforcement history of NPH action [21]. Within a task, hand usage is highly stable: healthy adults will sacrifice explicit task goals to maintain stable hand usage [22]; and most individuals continue to use their PH even after chronic injury leaves it less dexterous than their NPH [11]. However, less is known about psychosocial factors that influence hand usage in rehabilitation, because many assessments of function or hand use do not address the reasons and psychosocial aspects underlying that preference (for a review, see Bazo et al., 2023). Patients who must use their NPH after unilateral PH impairment may encounter psychosocial barriers to NPH use because hand usage is driven in part by history of success and confidence [8,24], Therefore, to help patients increase usage of their NPH after irreversible PH impairment, understanding the psychosocial aspects pertaining hand usage is important to better structure and prescribe tailored interventions. Click or tap here to enter text.

In the current study, we developed an inventory to measure psychosocial factors that could potentially influence hand usage. Specifically, we sought to identify participant experiences of comfort, concern, and fear related to use of their NPH – which, when identified, can be translated and applied to rehabilitation research. To this end, we first generated a large battery of sample items to measure four subjective (i.e. psychosocial) aspects of PH usage: self-reported PH performance, self-reported hand usage, concerns about physical consequences of NPH use, and concerns about emotional consequences of NPH use; and used an online service to collect responses from a large number of healthy individuals. We used the battery to identify a subset of items to assess those four categories in a shorter inventory, the Changed Hand Usage Concerns (CHUC).

## 2. METHODS

### 2.1. Study Overview

A large number of items were first generated based on clinical and scientific experience, to ensure relevance and comprehensiveness. An online web-based questionnaire (the “battery”) was then created to collect responses in a cross-sectional sample of healthy adults. Item responses were analyzed both psychometrically and clinically to identify a smaller set of items that would capture the underlying constructs among participant responses, and this smaller set of items comprises the CHUC inventory.

### 2.2. Participants

197 participants consented to participate in the study, via a consent sheet at the start of the survey that explained survey duration, data storage, principal investigator name, and study purpose. All participants who viewed the first page completed the first page (participation rate 100%). All participants were compensated for their time. Participants were excluded from analysis if they completed fewer than 90% of items (n=13) or failed both of two attention-check items (n=3), leading to a completion rate of 94% (200/213) and a final total of 181 analyzed participants. All further mention of participants refers to these 181.

Inclusion criteria were: US citizens age 18+ who self-reported as left handed or right handed (not ambidextrous) and used the Prolific survey distribution website (app.prolific.com) in December 2023, January 2024, or May 2025. The survey was open to any user of Prolific who met inclusion criteria. No advertisement or announcement was used; instead, potential participants were informed about the survey via their use of Prolific’s settings and notifications. Duplicate participants were prevented via Prolific’s unique IDs; no cookies or IP addresses were collected.

Participant ages were mean 38 ± std 12, median 36 (range 29-46); gender 82 male, 94 female, 2 other; self-reported handedness strongly left 21 (12%), moderately left 4 (2%), both hands equal 0, moderately right 23 (13%), strongly right 133 (73%). Full demographics are included in **Supplementary Table 1**. Handedness self-report was determined by the item “how would you describe your handedness?” Recruitment was not stratified by hand preference; the slight over-representation of left-handed participants (25/181; 14%) likely arose because left-handers were more likely to volunteer for a questionnaire related to handedness.

All procedures were approved by the Institutional Review Board of [institution removed for blinding]. No personal information was collected or stored. No statistical correction was applied to adjust for any non-representative aspects of the sample.

### 2.3. Comprehensive Item Battery

A large item battery was generated based on the scientific and clinical consensus of 3 subject matter experts (authors TS, AM, BAP), and tested for technical functionality before release. This battery was designed for comprehensiveness, and required approximately 15 minutes to complete. From this battery, the most useful items (as defined below) would be selected for inclusion in the final CHUC inventory. Participants were recruited via Prolific and data was collected via REDCap [25]. Battery questions were presented in fixed order to ensure structure and consistency.

The battery contained 138 items, divided into 4 categories (in order): Task-Specific (9 items), Concerns About Physical Limits (“Concerns-Physical,” 53 items), Actual-Use (24 items), and Concerns About Emotional Impacts (“Concerns-Emotional,” 52 items). Demographic items are not included in this list or analysis. All items used 5-point Likert-type scales, with response options and full item text shown in **Supplementary Table 2**. For example, one item was “How do you think your overall comfort with using your left hand compares to other righties?,” with the following options: Much Worse, Somewhat Worse, About Average, Somewhat Better, and Much Better. Within each item, left/right labels were adjusted according to the individual’s self-reported hand preference.

Critically, all four categories reflect subjective factors. Task-Specific reflects participants’ (psychosocial) belief about task-specific NPH function. Concerns-Physical reflects their (psychosocial) concern about physical consequences of NPH use, Actual-Use reflects their (psychosocial) self-reported beliefs about NPH use. Concerns-Emotional reflects their (psychosocial) concern about emotional consequences of NPH use.

The Task-Specific category comprised 9 items; one on self-reported hand preference, followed by 8 items: 4 Topics (overall comfort with NPH, NPH drawing accuracy, NPH drawing speed, likelihood of catching tennis ball with NPH), each with two levels of Objectivity (objective, relative to other people of the same hand preference). The “Topics” were selected to capture activities that have a PH advantage but different task requirements (drawing = fine motor, closed environmental conditions; catching = gross motor, open environmental conditions), because these categories of motor tasks (fine vs. gross and closed vs. open) can influence hand preference [26].

The other three categories (Concerns-Physical, Actual-Use, and Concerns-Emotional) all used the list of 13 activities shown in **Table 1**. These activities were selected to include activities of daily living (e.g. dressing, washing), instrumental activities of daily living (e.g. cooking, shopping), social activity (e.g. eating in a restaurant, playing cards), stabilization (e.g. carrying a tray), bimanual activities (e.g. opening a jar), unimanual activities (e.g. using a fork), and activities outside the home (e.g. shopping, restaurant).

**Table 1:**
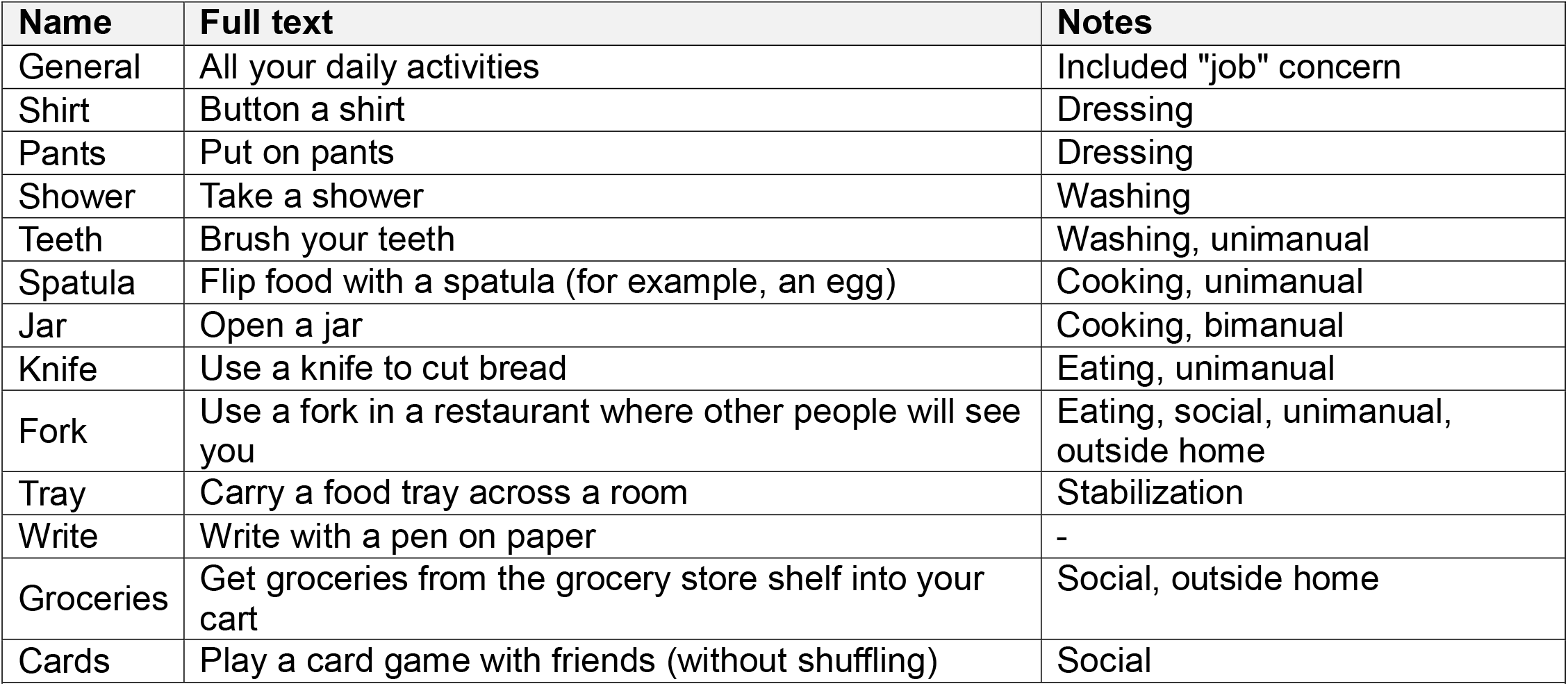
Activities shared across the Concerns-Physical, Actual-Use, and Concerns-Emotional categories of. Activities are listed as “unimanual” or “bimanual” if they fit unambiguously into one of those descriptions. For example, “Write” is listed as neither because it has a bimanual component (paper stabilization) but most people consider it unimanual.

| Name | Full text | Notes |
| --- | --- | --- |
| General | All your daily activities | Included "job" concern |
| Shirt | Button a shirt | Dressing |
| Pants | Put on pants | Dressing |
| Shower | Take a shower | Washing |
| Teeth | Brush your teeth | Washing, unimanual |
| Spatula | Flip food with a spatula (for example, an egg) | Cooking, unimanual |
| Jar | Open a jar | Cooking, bimanual |
| Knife | Use a knife to cut bread | Eating, unimanual |
| Fork | Use a fork in a restaurant where other people will see you | Eating, social, unimanual, outside home |
| Tray | Carry a food tray across a room | Stabilization |
| Write | Write with a pen on paper | - |
| Groceries | Get groceries from the grocery store shelf into your cart | Social, outside home |
| Cards | Play a card game with friends (without shuffling) | Social |

In the Concerns-Physical category, participants were told to imagine they had to favor their NPH because their PH was injured (unable to move or separate their fingers) and asked how concerned they would be about 4 concerns on each activity: success, time, habits, and availability of help, as shown in **Figure 1A**. In the “all daily activities” item, participants were asked about one additional concern: “whether you could perform well at your job.”

**Figure 1:**
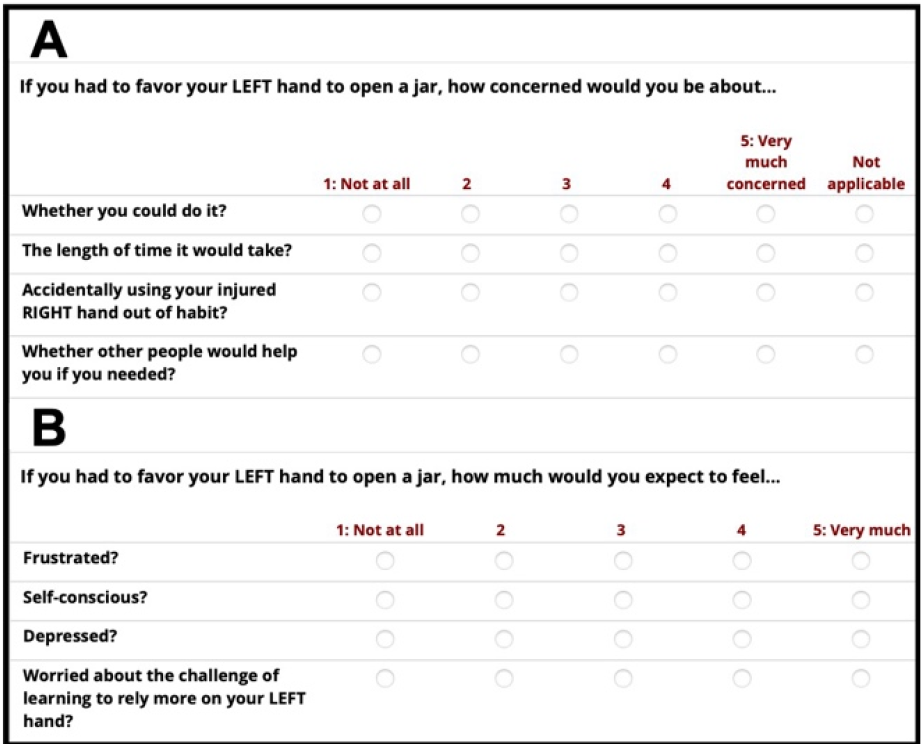
Sample battery items for one activity in a right-handed participant, showing **(A)** concerns-physical and **(B)** concerns-emotional. Both categories were preceded with the context, “Imagine that you had to favor your LEFT hand because your RIGHT hand was injured (unable to separate or move your fingers).”

In the Actual-Use category, for each activity, participants were asked whether they use one hand or two. Based on their response, the questionnaire included one of two possible follow-up items: If they used one hand, they were asked which hand (always NPH, usually NPH, either hand equally, usually PH, always PH); if they used both hands, they were asked how often they use their NPH (very rarely, somewhat rarely, about half the time, somewhat often, or very often). For analyses, the two follow-up items were merged into a single “how often do you use your NPH?” item. The “all daily activities” item was omitted from the Actual-Use category.

In the Concerns-Emotional category, participants were told to imagine they had to favor their NPH because their PH was injured (unable to move or separate their fingers) and asked whether they would expect to feel 4 concerns: frustrated, self-conscious, depressed, or worried about the challenge, as shown in **Figure 1B**. Participants also answered 4 additional items: the same 4 concerns in a general “all activities” context.

Critically, all questions asked participants to report their concern about potential consequences. Therefore, even for items about physical limitations and actual use, the battery’s nature as a self-report inventory means its answers reflect subjective (psychosocial) characteristics.

Participants did receive the opportunity to review their answers before finalizing their answers.

### 2.4. Data Analysis

To describe how participant responses depended on activities and other factors, a series of ANOVAs were performed on mean Likert-type scores. ANOVAs were performed in MATLAB 23.2.0 using the ‘anovan’ function. For the categories Physical Concerns and Emotional Concerns, the ANOVAs were 12 (Activity) * 4 (Concern); for the category Task-Specific, 4 (Topic: general comfort, drawing accuracy, drawing speed, tennis) * 2 (Objectivity); and for the category Actual-Use, a one-way (Activity; 12 levels) ANOVA. In all cases, the “general activities” items were omitted from analysis. Data with missing values were omitted from analysis; in Concerns-Physical missing values were 52/8688 samples (0.06%) and zero samples in all other categories. Post-hoc tests were performed using Tukey’s Honestly Significant Difference.

Subsequently, a five-step process was used to develop the CHUC inventory from the battery responses:

First, the categories were assessed for internal consistency via reliability analysis, performed in SPSS 29 (IBM, Armonk NY) by calculating Cronbach alpha, separately for each category. If removing an item would substantially increase the category’s reliability (by more than 0.01), the item was flagged for potential removal and the reliability analysis was performed without that item. Second, latent factors in the data were identified via PCA, performed in MATLAB version 23.2.0 (Mathworks, Natick MA). Third, the factor loadings (contributions of each item to the top latent factors) were obtained. The top factors were identified within each category as the largest factors that together explained ≥80% of the variance, and then by removing any factors with eigenvalue < 1. Fourth, to reduce the number of items, items were removed from the inventory based on the analyses. Authors TS and BAP removed items based on the following criteria: ill-fitting items (flagged for removal in reliability analysis), non-discriminative or weakly loading items (relatively low absolute loadings on all factors), redundant items (followed the same pattern of absolute loadings as other items that had higher absolute loadings and/or loaded on more factors), separable items (an item that loads primarily on a factor with no other strongly loading item), and inventory-consistency requirements (if a concern is removed, it must be removed for all activities within the category; if an activity is removed, it must be removed for all categories).

Fifth and last, to confirm that the item reduction did not compromise internal consistency, the reliability analysis was repeated for the reduced set of items (i.e. the items that comprise CHUC).

## 3. RESULTS

### 3.1. Descriptive statistics on participant responses

Participants completed the battery in 15.8 (8.3) minutes (minimum 5.8, max 51.1, median 13.5). Overall descriptive results are shown in **Figure 2**. We explored the pattern of effects by performing an ANOVA within each category, with results detailed in the following paragraphs.

**Figure 2:**
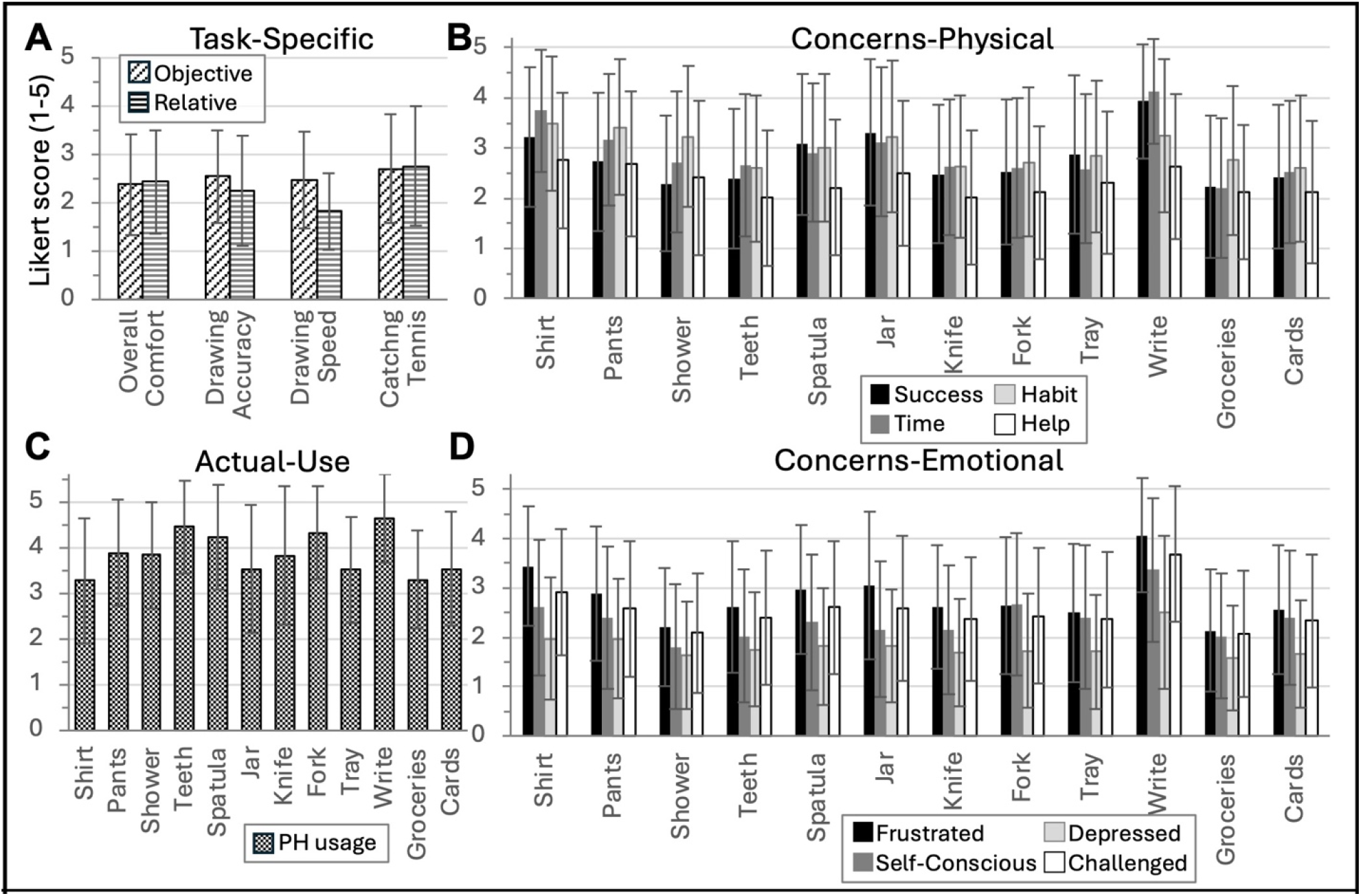
Descriptive statistics for battery responses, mean (SD). PH = preferred hand. **(A)** Task-Specific category. **(B)** Concerns-Physical category. **(C)** Actual-Use category. **(D)** Concerns-Emotional category.

In the Task-Specific category (**Figure 2A**), we found significant effects of Topic (F_3,1440_ = 18.7, p = 7.0×10^-12^), Objective (F_1,1440_ = 14.0, p = 1.9×10^-4^), and Interaction (F_3,1440_ = 9.2, p = 5.0×10^-6^). Post-hoc tests showed that the topic effect was driven by people scoring Drawing Speed lower than other topics, and Tennis higher than other topics; that the Objectivity effect represented people scoring their relative performance (i.e. compared to others) higher than their objective performance, and that the interaction effect arose from a stronger Objectivity effect for Drawing Speed.

In the Concerns-Physical category (**Figure 2B**), we found significant effects of Activity (F_11,8588_ = 52.3, p = 2.0×10^-112^), Concern (F_3,8588_ = 96.3, p = 2.8×10^-61^), and their interaction (F_33,8588_ = 5.1, p = 4.7×10^-20^). Post-hoc tests revealed that the Activity effect included differences between 43/66 pairwise comparisons; full results are shown in **Supplementary Table 3** and summarized here by ranking the activities from highest to lowest average physical concern: Write, Shirt, Jar, Pants, Spatula, Tray, Shower, Fork, Knife, Cards, Teeth, Groceries. The Concern effect was driven by lower concern for the availability of help than any other concern, and lower concern for success than time or habit; and the interaction effect included significant differences in 436/1128 comparisons (**Supplementary Table 4)**. Overall, concerns about physical limits depended greatly on the specifics of activity and concern.

In the Actual-Use category (**Figure 2C**) we found a significant effect of its only factor, Activity (F_11,2160_ = 26.4, p = 4.7×10^-52^). This effect involved significant differences in 35/67 comparisons, detailed in **Supplementary Table 5** and summarized by ranking the activities from highest to lowest PH use as: Write, Teeth, Fork, Spatula, Pants, Shower, Knife, Jar, Cards, Tray, Groceries, Shirt.

In the Concerns-Emotional category (**Figure 2D**) we found significant effects of Activity (F_11,8640_ = 65.2, p = 1.1×10^-140^), Concern (F_3,8640_ = 221.9, p = 1.1×10^-138^), and their interaction (F_33,8640_ = 3.2, p = 2.8×10^-9^). Post-hoc tests revealed that the Activity effect included differences between 39/66 pairs (**Supplementary Table 6**), and the ranking from highest to lowest average emotional concern was: write, shirt, pants, spatula, jar, fork, cards, tray, knife, teeth, groceries, shower. For the Concern effect, all pairings were significantly different; the ranking from strongest to weakest was Frustrated, Challenged, Self-Conscious, Depressed. The interaction effect included significant differences in 515/1128 comparisons (**Supplementary Table 7**).

For an overall comparison of Concerns-Physical vs. -Emotional (averaging across concerns in each), the two categories were correlated across activities (Spearman ρ = 0.783 p = 0.002). The same activities led to strongest expectations of both Physical and Emotional Concern: the highest-scored activities in both were write, shirt, and then (in different orders) pants, spatula, and jar; though overall the ranking of activities was not correlated between Emotional vs Physical (Kendall’s *τ* = 0.03, p = 0.947). Therefore, activity choice is a major factor – but not the sole factor – in determining overall concern about the consequences of NPH use.

### 3.2. Item categories were internally consistent

To maximize internal consistency within each category, we performed a reliability analysis (Cronbach’s *α*). First, we performed a reliability analysis on all categories in the “original battery” (**Table 2** column 1), omitting only the binary (yes/no) items in the Actual-Use category (i.e. “do you use one hand or two?”). This analysis revealed two items that could be removed to increase reliability: TaskSpecific-HandednessSelfDescribe and ActualUse-Write. We removed these items and performed a reliability analysis on the remaining “filtered battery” (**Table 2** column 2), in which all categories achieved high internal consistency (□ > 0.7) [27].

**Table 2.** Reliability analyses. Values are Cronbach’s α. After filtering, all major categories achieved high internal consistency (α > 0.7), 3 of which remained after item reduction (equivalent to the CHUC).

| Category | Original battery | Filtered battery | After item reduction (=CHUC) |
| --- | --- | --- | --- |
| Task-Specific | 0.725 (9 items) | 0.803 (8 items) | 0.751 (6 items) |
| Concerns-Physical | 0.970 (53 items) | 0.970 (53 items) | 0.920 (21 items) |
| Actual-Use | 0.729 (12 items) | 0.731 (11 items) | 0.536 (7 items) |
| Concerns-Emotional | 0.977 (52 items) | 0.977 (52 items) | 0.929 (14 items) |

### 3.3. Principal components analysis revealed 3-13 latent factors per category

We used PCA to identify latent factors in the filtered battery, which revealed that each category had 3-13 factors that together explained 80% of the variability, while omitting any factors of eigenvalue < 1 (**Figure 3)**. For all categories, the first component accounted for a plurality of variance (30-39%); this first component always represented covariance between all items within a category, as shown in the full set of factor loadings (**Supplementary Tables 8-11)**. These 3-13 latent factors were used to perform item reduction as discussed in the following section.

**Figure 3:**
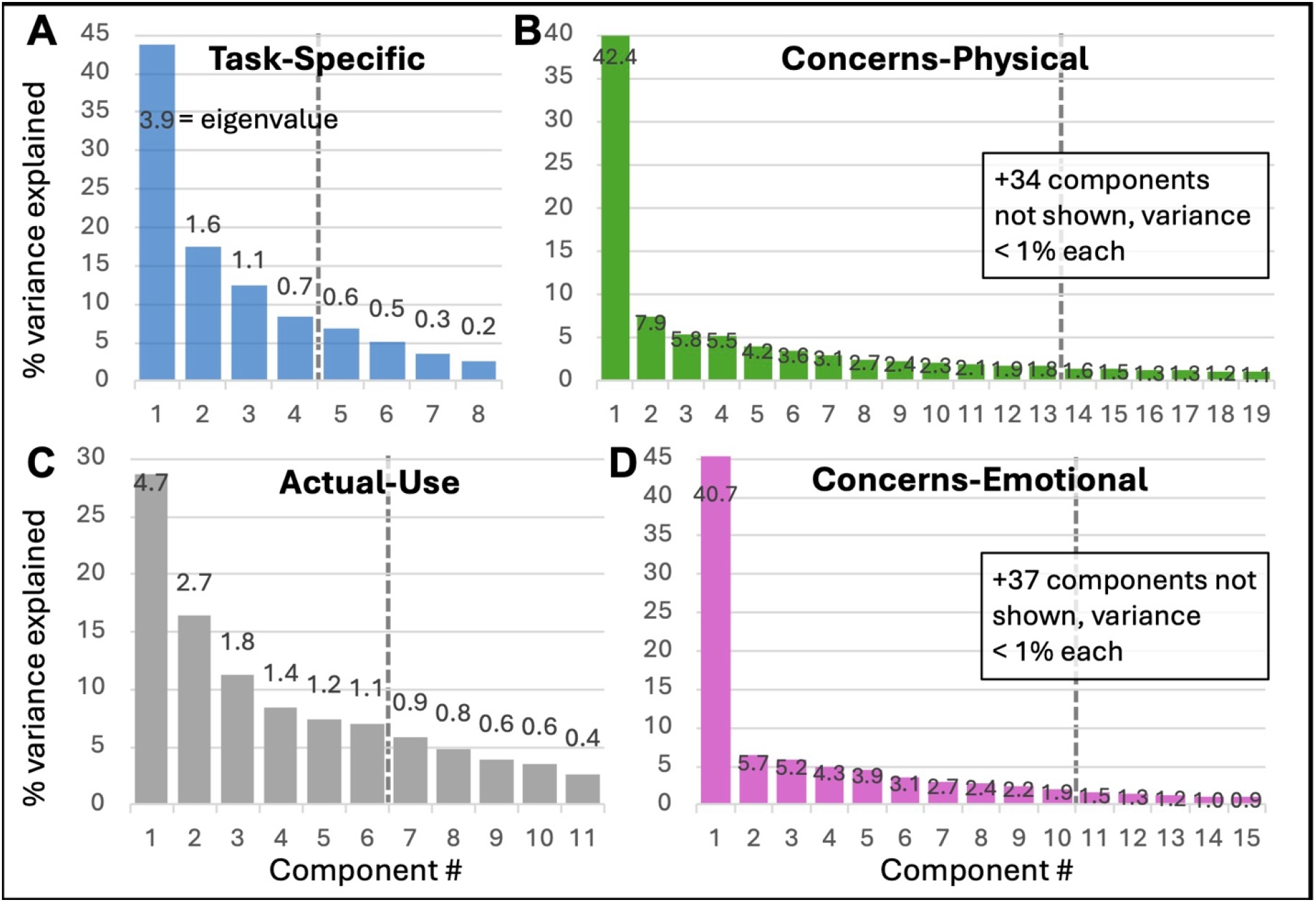
Scree plots of latent factors from PCA of question battery. Eigenvalue listed at each bar. Factors were reviewed for item reduction that together accounted for 80% of variance (dashed line), omitting factors with eigenvalue (numbers at each bar) < 1.0. **(A)** Task-Specific category. **(B)** Concerns-Physical category. **(C)** Actual Use category. **(D)** Concerns-Emotional category.

### 3.4. Item reduction removed 90/138 battery items

To reduce the size of the battery, 90 items were removed out of the original 138, following the decisions listed in **Table 3**. An objective of this study was to reduce the battery size (i.e. to create a small feasible final inventory to reduce respondent burden without sacrificing psychometric properties), so we applied the exclusion criteria rigorously and aggressively to achieve the item-reduction goal.

**Table 3.** Item reduction based on results of PCA. * = wildcard, all matching items.

| Category | Item(s) removed | Reason |
| --- | --- | --- |
| Task-Specific | HandednessSelfDescribe | Ill-fitting (removed in reliability analysis) |
|  | Draw-fast-* | Redundant with Draw-acc-* |
| Concerns-Physical | Physical-general-* | Weak |
|  | Physical-*-time | Redundant with *-succeed |
|  | Physical-pants-* | Weak |
|  | Physical-shower-* | Redundant with teeth-* & shirt-* |
|  | Physical-spatula-* | Redundant with write-* & jar-* |
|  | Physical-knife-* | Weak |
|  | Physical-groceries-* | Internal consistency (Emotional-groceries removed) |
| Actual-Use | Actual-pants | Internal consistency (Physical-pants removed) |
|  | Actual-shower | Internal consistency (Physical-shower removed) |
|  | Actual-spatula | Internal consistency (Physical-spatula removed) |
|  | Actual-knife | Internal consistency (Physical-knife removed) |
|  | Actual-groceries | Internal consistency (Emotional-groceries removed) |
| Concerns-Emotional | Emotional-general-* | Internal consistency (Physical-general removed) |
|  | Emotional-*-selfconsc | Redundant with Emotional-*-depressed |
|  | Emotional-*-challenged | Redundant with Emotional-*-frustrated |
|  | Emotional-pants-* | Internal consistency (Physical-pants removed) |
|  | Emotional-shower-* | Internal consistency (Physical-shower removed) |
|  | Emotional-spatula-* | Internal consistency (Physical-spatula removed) |
|  | Emotional-knife-* | Internal consistency (Physical-knife removed) |
|  | Emotional-groceries-* | Weak |

“ActualUse-Write” was readmitted in analysis at this stage, even though it was ill-fitting (its removal increased reliability, as described above). We retained ActualUse-Writing because writing is a primary predictor of hand preference [28].

Notably, items that included a social element were more likely to survive the item reduction process. The physical concern “availability of help” was relatively independent of other physical concerns; and comparison between “fork” vs “knife” activities provide a key illustration, because they are similar activities (utensils) but distinguished in the battery via the social context we added to the “fork” activity. The two activities had redundant loadings on the latent factors of physical concerns – but absolute loadings across the 12 latent factors were higher for “fork” (mean 0.118 (0.14)) than “knife” (mean 0.101 (0.12)). In other words, for two related and redundant activities, the one with the social aspect (“fork”) explained more of the variability in responses.

To evaluate our item reduction efforts, we performed a reliability analysis on those retained items after item reduction (**Table 2** column 3). The category Actual-Use was no longer internally consistent (*α* = 0.536), but the primary categories Concerns-Physical and Concerns-Emotional remained highly internally consistent (*α* ≥ 0.92) despite our rigorous and aggressive item reduction.

### 3.5. Final Inventory Development

The CHUC inventory was defined as the 51 items shown in **Figure 4**. This included 7 task-specific items + [7 activities (shirt, teeth, jar, fork, tray, write, cards) * 3 concerns-physical (succeed, habit, help) * 1 actual-use * 2 concerns-emotional (frustration, self-conscious)] + 3 narrative items.

**Figure 4A:**
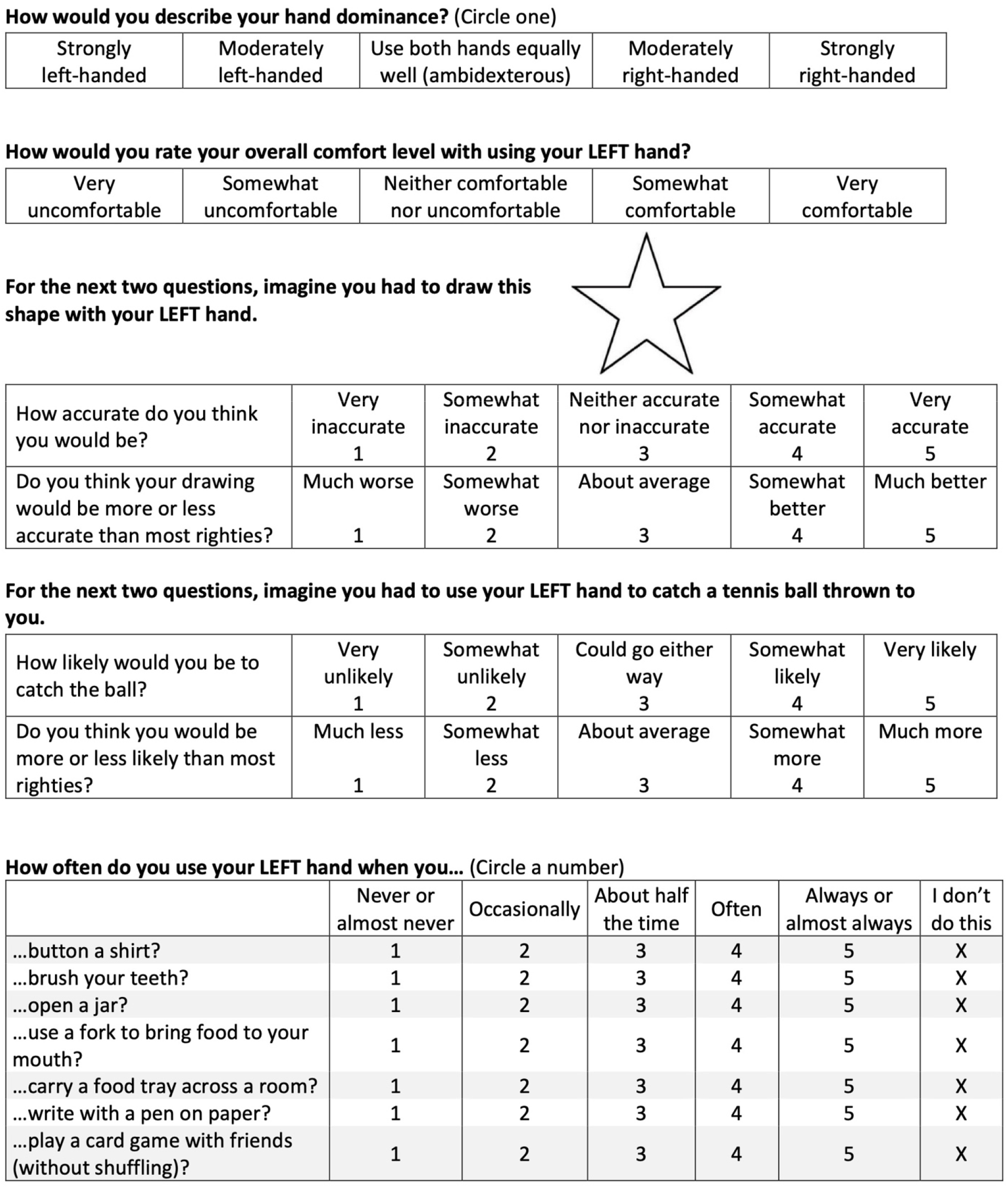
CHUC inventory (right-handed healthy participant), page 1/3.

**Figure 4B:**
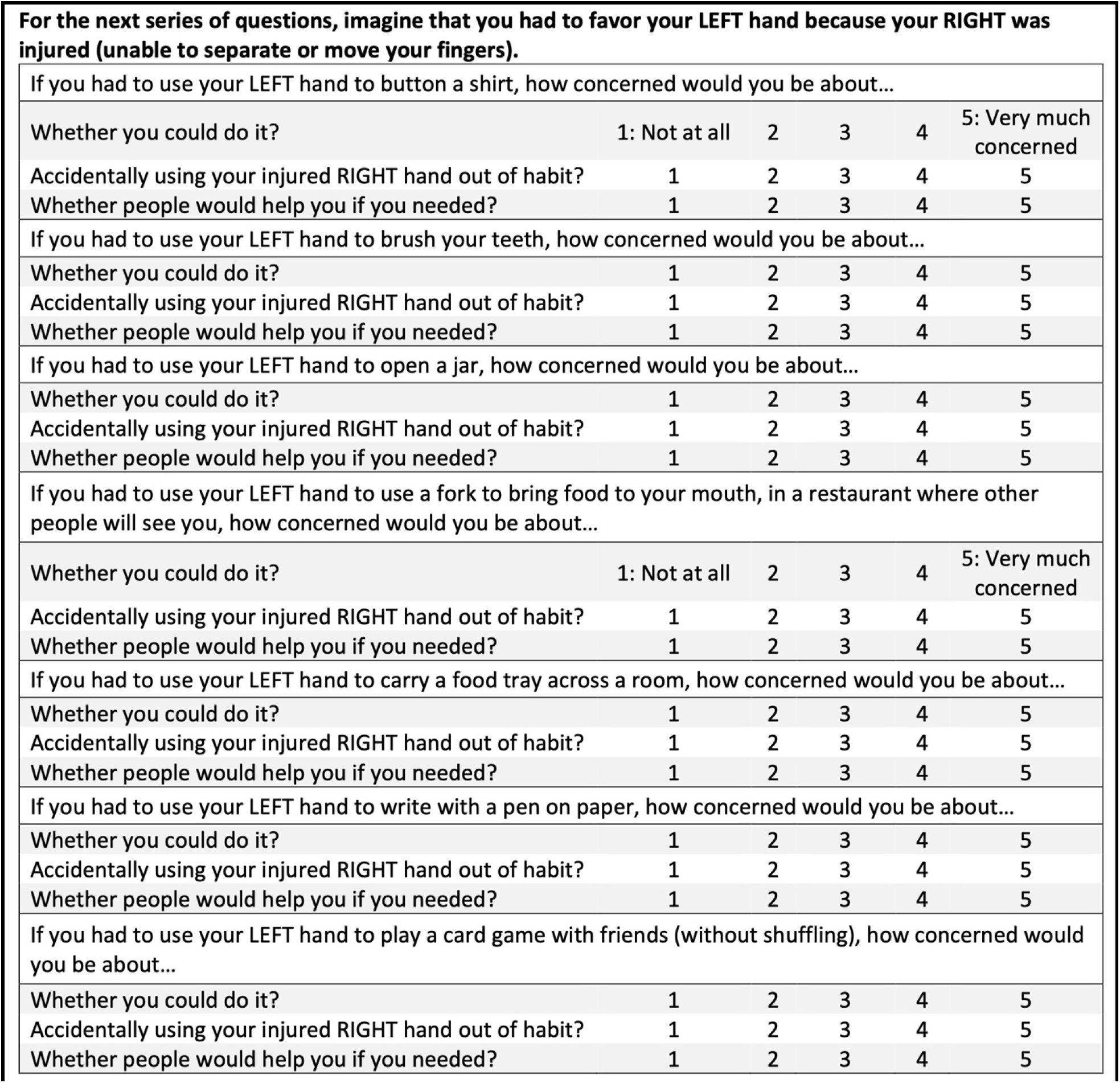
CHUC inventory (right-handed healthy participant), page 2/3

**Figure 4C:**
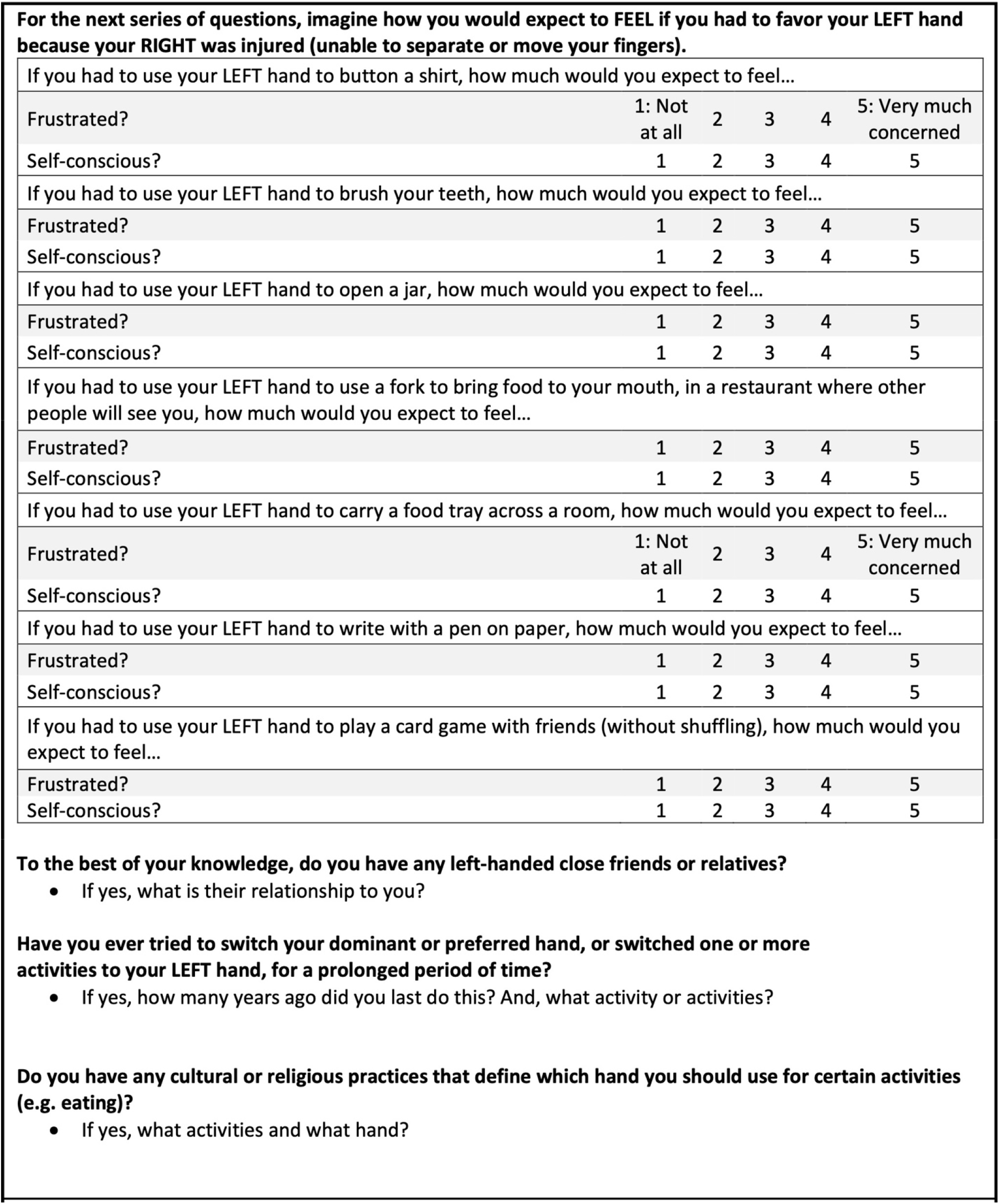
CHUC inventory (right-handed healthy participant), page 3/3

The CHUC’s narrative items capture additional factors that could influence emotional reactions to hand choice (family history, personal history, and cultural/religious history) and are detailed at the bottom of **Figure 4C**.

The item “hand preference self-description” had been removed in earlier steps of the current analysis but was reintroduced to the CHUC (included among the 7 task-specific items) because the authors considered it necessary for the study of hand choices.

The CHUC includes two additional changes to accommodate the option of paper-based implementation. First, whereas the battery’s Actual-Use category addressed each activity with two items using branching logic, the CHUC presents each Actual-Use activity via a single item. Second, we reordered the categories (to: Task-Specific, Actual-Use, Concerns-Physical, then Concerns-Emotional) to prevent page breaks within categories.

## 4. DISCUSSION

We tested an online battery of 138 items and reduced it to the 51-item (3 page) “Changed Hand Usage Concerns” (CHUC) inventory. We found that “concerns about physical challenges of using the NPH” and “concerns about emotional challenges of using the NPH” reflect coherent internally-consistent constructs, which are partly but not completely correlated. Activities and concerns with social aspects often had strong contributions to participant responses. The CHUC inventory provides a brief assessment of psychosocial factors that may influence hand usage, which can now be translated and applied to rehabilitation research with patients who have – or should – change their hand usage after unilateral impairment [10].

### 4.1. Implications for Rehabilitation

Hand choice assessment is important for rehabilitation because patients may change their hand choice patterns after asymmetric impairment such as stroke [29,30] and peripheral nerve injury [22,31]. These altered choice patterns impact patient outcomes: use of the impaired hand – independently from function of the impaired hand – is associated with patient quality of life, at least after peripheral nerve injury [10]. However, the causes of hand choice are not well-understood because those causes are not limited to physical factors [11,17,21,30]. As a result, no reliable methods exist to promote the use of one hand over another [13]. CHUC provides the first tool to assess the non-physical (i.e. psychosocial) factors that may influence hand choice, and thereby provides a necessary first step toward manipulating those psychosocial factors and promoting patient-appropriate hand choices.

### 4.2. The CHUC inventory has feasibility, content validity, and initial reliability

Our primary goal was to create an inventory short enough to include in clinical care or research, while still capturing the key latent factors that explain participant responses. The final CHUC inventory contains 51 items, covering three sheets of paper; all items are 5-point Likert-type scales, except for three optional narrative items at the end. We estimate that the CHUC should take under 5 minutes to complete because it contains 37% as many items as the battery (which had median duration of 12.7 minutes).

The current study established 2-3 forms of validity for CHUC [32]. We established content validity by systematically determining the item contents, factorial validity by selecting items based on principal components analysis and demonstrating the relationship between facets (Concerns-Physical vs. -Emotional, see next section); and partly established reliability by demonstrating the internal consistency of its constructs. Other psychometric properties (e.g. stability, sensibility, applicability, factorial validity) should be established in a patient population, now that CHUC has been created. Nevertheless, CHUC’s 2 established psychometric properties make it as well-validated as 12/13 existing lateral preference inventories, which have established a median of 0 psychometric properties per inventory (range 0-3) [33].

### 4.3. CHUC measures consistent groups of psychosocial factors: concerns about physical limits and emotional impacts

Our two main categories of items, Concerns-Physical and Concerns-Emotional, had extremely high internal consistency (*α* ≥ 0.92), suggesting that each of them may represent a singular underlying construct.

While we describe our categories as Concerns-Physical and Concerns-Emotional, the battery (and CHUC) record participant self-reports, so all answers reflect subjective psychosocial factors. Concerns-Physical reflects their (psychosocial) concern about physical consequences of NPH use, and Concerns-Emotional reflects their (psychosocial) concern about emotional consequences of NPH use. Similarly, the category Actual-Use reflects self-report and thus self-perceived use of the NPH, which may differ from objective measures of NPH use. The Actual-Use category had poor internal consistency across items (topics/activities), which matches our descriptive result that self-reported NPH use depends on the activity (**Figure 2C**), just as does actual hand use [19].

Because the CHUC measures concerns about future/potential changes, it may provide useful information when completed by patients about to begin rehabilitation periods requiring a handedness shift. Specifically, CHUC results may allow professionals to tailor and facilitate the intervention. Additionally, researchers could use the CHUC to select, stratify, or analyze participants in research studies of handedness shifts. Large-scale application of the CHUC may lead to better understanding of why humans rarely succeed at shifting handedness [13].

### 4.4. Analysis of the question battery reveals partial similarity between emotional and physical concerns

We described the full results of our battery questionnaire, to provide descriptive data on psychosocial concerns about activities. We found a significant but moderate correlation between concerns about physical limits vs. concerns about emotional consequences, and that concern levels were always activity-dependent, with global highest concerns about writing and putting on a shirt. While ‘amount of concern’ was not a criterion in our item reduction process, those two highest-concern activities were both included in CHUC.

### 4.5. Limitations & Future Directions

To create the brief feasible CHUC inventory, we had to begin with a battery questionnaire that was too long for easy delivery, and collect a large sample of participants despite the battery’s length. Therefore, we used an online study recruitment tool (Prolific). As a result, it was not possible to collect data on the participants’ actual hand use. Now that the CHUC is available for in-person research, future studies can determine further psychometric properties of CHUC, including its applicability: i.e. whether and how CHUC results correspond to actual hand usage and performance.

All participants in the current study were healthy adults. For delivering the CHUC to patients with relevant physical impairments, we recommend adding 1-2 additional items: First, “How confident are you that you will recover from [your impairment]?” would be a 5-point Likert-type with response options from “Not at all confident” to “very confident.” Second, if appropriate for the patient group, “Which side or sides of your body are affected by [your impairment]?” would be a 5-point Likert-type with response options from “Entirely left” to “Entirely right.”

## 5. CONCLUSIONS

The Changed Hand Usage Concerns (CHUC) inventory provides the first assessment of psychosocial factors that may influence hand use. CHUC is brief, partially validated, measures internally consistent categories (psychosocial concerns about physical & emotional consequences of non-preferred hand usage), and includes activities with social aspects. CHUC will allow future research to identify how these psychosocial factors correspond to actual measurements of hand use in healthy adults and patients with unilateral impairment.

## Data Availability

All data are available online at the Open Science Framework repository

https:/doi.org/10.17605/OSF.IO/3UF2C

## Abbreviation List

CHUC: Changed Hand Usage Concerns inventory
PH: Preferred hand
NPH: Non-preferred hand
PCA: Principal Components Analysis

## Acknowledgments

The authors would like to thank Elizabeth Woollen for her work creating an initial pilot version of the CHUC, and Brian Johnson for his help with online survey methods.

